# Bacterial Contamination of Inanimate Surfaces and Equipment, Distribution and Susceptibility Patterns in Pediatric Wards at a Northern Tanzania Zonal Referral Hospital

**DOI:** 10.1101/2025.07.08.25331103

**Authors:** Zacharia L Laizer, Imelda N Philbert, Anthony S Charles, Kelvin Musa, Jovin R Tibendarana, Patrick Lucas Mabula, Reuben S. Maghembe, Aisa Shayo, Sixbert I Mkumbaye, Nancy A Kassam, Debora Charles Kajeguka

**Author notes:** Corresponding Author: Zacharia L Laizer. **Other Authors E-mails** Imelda N Philbert Anthony S Charles Kelvin M Musa, Jovin R Tibenderana, Reuben S. Maghembe Aisa Shayo, Patrick Mabula Sixbert I Mkumbaye Nancy A Kassam, Debora Charles Kajeguka.

## Abstract

**Background:** Hospital-acquired infections (HAIs) represent a significant global health concern, contributing to high rates of morbidity and mortality. Globally, bacterial contamination of hospital surfaces and equipment has been identified as a critical pathway for the transmission of nosocomial infections. Despite its significance, the extent of surface and equipment contamination in pediatric wards at Kilimanjaro Christian Medical Centre (KCMC) remains underexplored. This study aims to determine the proportion of bacterial contamination of inanimate surfaces and equipment, distribution and susceptibility patterns in pediatric wards at Northern Tanzania Zonal Referral Hospital.

**Methodology:** A descriptive cross-sectional hospital-based study conducted in pediatric wards from May to August 2021. The study was conducted at Kilimanjaro Christian Medical Center, a Northern Tanzania Zonal referral Hospital. Swabs were collected from inanimate surfaces and equipment and cultured on general-purpose media, differential media, and enriched media which were MacConkey agar, Blood agar and Chocolate agar to identify the bacteria isolated. Data was entered and analyzed using Statistical Package for the Social Sciences (SPSS).

**Results:** 206 (86.6%) were positive for bacterial growth. Gram-negative bacteria were the most predominant bacteria identified 304 (82.4%) in which *E. coli* was the leading isolate 114 (30.9%), followed by *Pseudomonas aeruginosa* 93 (25.2%), *Klebsiella oxytoca* 44 (11.9%), *Klebsiella pneumonia* 41 (11.1%), *Acinetobacter species* 10 (2.7%), *Proteus vulgaris* 1 (0.3%) and *Citrobacter* species 1 (0.3%). Gram-positive isolates were 65 (17.6%) in which *CoNS* were 58 (15.7%) and *S. aureus* 7 (1.9%).

**Conclusion:** In pediatric wards, surfaces and equipment can harbor diverse pathogenic bacteria with the potential to cause hospital-acquired infections to patients especially immuno-compromised or critically ill patients. To mitigate this risk, it is crucial to assess infection prevention practices, implement regular disinfection programs and raise awareness among healthcare professionals about the potential for pathogen transmission from hospital surfaces and equipment.

## Introduction

Bacterial contamination refers to biological contamination whereby bacteria spread via physical means (sharing media and reagents using unplugged pipettes and abnormal contact with inanimate objects) and biological means (direct or indirect contact on hands) ^1^. Bacterial contamination may be of equipment for direct patient care (Stethoscope and Ultrasound equipment) and object surfaces that are used in clinical data and management such as medical charts, computer keyboards and mobile phones ^2^. There is increasing evidence that contaminated inanimate surfaces especially those frequently touched with bare hands can contribute to the spread of healthcare-associated pathogens ^3^.

Hospital-acquired infections (HAIs) or nosocomial infections (NIs) are of huge concern around the world and account for the most common deaths in hospitalized patients, increasing the cost of health care system ^4^^5^.

Various studies conducted worldwide have reported different prevalence rates of HAIs. Most of these studies identified *Escherichia coli, Staphylococcus aureus, Klebsiella species,* and *coagulase-negative staphylococci* as the leading pathogens associated with HAIs and antimicrobial resistance ^6–9^.

In Sub-Saharan Africa, several systematic review analyses have been conducted to assess the burden HAIs, reporting a variety of microorganisms. Among the most frequently isolated were *Escherichia coli, Pseudomonas aeruginosa, Acinetobacter baumannii,* and *Citrobacter species.* The prevalence of HAIs was analyzed by subregions wich were West Africa, Southern Africa, East Africa, and Central Africa with a pooled prevalence estimated between 12.7% and 12.9%^10–12^.

According to National Infection Prevention and Control Guidelines for Health Care Services in Tanzania ^13^, these infections arise during the course of medical or surgical treatment and are often linked to poor sanitation practices, inadequate decontamination procedures, prolonged use of invasive devices and the overuse of antibiotics, which fosters antimicrobial resistance (AMR). In Tanzania, it is estimated that 15 out of every 100 patients receiving care acquire an HAI ^14^. Common pathogens responsible for HAIs include *Escherichia coli, Pseudomonas aeruginosa, Klebsiella* species, and *Staphylococcus aureus*, all of which are prevalent in this study’s findings.

Various reports have pointed to the role of healthcare workers in the transmission of bacterial pathogens to patients, pathogens may be transmitted directly from contaminated surface to susceptible patients ^15,16^. A study done in Baghdad hospital reported that most of medical staffs are the major contaminants where they move back and forth between operating rooms and other parts of hospital wards without changing their gowns and slippers ^17^. A systematic review reported that surface contaminants in operating procedures were mostly originated from staffs where by opportunistic pathogens associated with skin of personnel causes more than half of the bacterial infection after clean surgery ^18^. Excessive contact with computer keyboards and carelessness of personnel led to contamination hence source of NIs ^5^. NIs affects children more due to their nature of weak immune system. Poor hygienic measures in hospitals and lack of proper management of the patients further aggravate the situation hence cause complications, such as; antibiotic resistance, patient morbidity and mortality ^19^.

On the other hand, several studies have reported evidence of contamination of some commonly used equipment in hospital wards, the most frequently identified pathogens were Gram positives such as *Staphylococcus aureus*, *Vancomycin Resistant Enterococci* (VRE), *Coagulase negative staphylococcus, Methicillin Resistant Staphylococcus aureus* and gram negatives were *Pseudomonas aeruginosa, non-fermenting gram-negative bacteria*, *Acinetobacter baumannii* and *Klebsiella species* ^20^^21 2 3^.

In Uganda, a study reported that among all the equipment and surfaces sampled 44.2% of them presented with bacterial growth, the common bacteria isolated were Gram positive 75.4%of which the most isolated was *Staphylococcus aureus*, whereas 24.6% were Gram negatives that consisted of *Klebsiella pneumoniae*, *Proteus vulgaris*, *Enterobacter species* and *Serratia mercescens* ^22^. Triggered by all these lines of evidence, this work explored the risk of pathogenic bacterial contaminations with antimicrobial resistance profiles in pediatric wards at Kilimanjaro Christian Medical Centre, a Northern Tanzania zonal referral Hospital. Here we report a versatile pattern of diverse bacterial pathogens posing risks of nosocomial infection with MDR cases.

Despite the studies done in Africa and East Africa, in Tanzania, there is no published information on the bacterial contamination of inanimate surfaces and equipment, distribution and susceptibility patterns in pediatric wards. Therefore, this study aims to determine bacterial contamination of inanimate surfaces and equipment, distribution and susceptibility patterns in pediatric wards at Northern Tanzania Zonal Hospital where data to be obtained will notify the National Infection Prevention and Control Standards for Hospitals in Tanzania to contribute and deliver safe health care and social services in pediatric wards, also the study will put much prominence on best practices in infection prevention and control to prevent bacterial contaminations which cause NIs, particularly in pediatric wards.

## Methodology

### Study Area

The study was conducted at KCMC Hospital located in Moshi-Kilimanjaro, Tanzania. KCMC is a Northern Zone referral Hospital serving approximately eleven million people 11,000,000 in Northern Tanzania as per the 2022 census. The pediatric ward is divided into four sections which are Pediatric One (P1), Pediatric Two (P2), Pediatric Three (P3) and Pediatric Intensive Care Unit (PICU). Pediatric one consists of children with the age of more than one month, Pediatric two consists of children with Surgical and Urological cases, Pediatric three consists of children with the age of 1 hour to one month and Pediatric Intensive Care Unit consists of children with serious cases.

### Study Design

This was a descriptive cross-sectional hospital-based study conducted from May 2021 to August 2021 at Kilimanjaro Christian Medical Center (KCMC) in pediatric wards. The study population was inanimate surfaces and equipment found in pediatric wards in which the purposive sampling technique was employed, where frequently used and touched equipment and inanimate surfaces were included in the study. Inanimate surfaces and equipment found in pediatric wards which were considered to be the source of infection were included in the study. Such equipment were Stethoscopes, Thermometers, Mechanical ventilators and Electronic equipment and inanimate surfaces were, Baby coats, Washing sink taps, Door handles, Bedside tables, Patient beds and Medical Charts. Bed sheets, latrines and sharp boxes were excluded in the study. The sample size was determined using the Fisher’s formula n=Z^2^ P(1-P)/E^2^ Where, n – Number of the minimum sample size required, Z–Level of confidence (1.96 for 95% confidence interval) P-Previous prevalence (19.26% prevalence of bacterial contamination done in pediatric wards in Xapala city, Mexico)^23^ and E-Absolute error (5%)

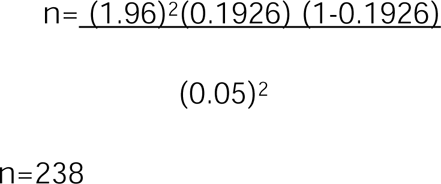

The estimated sample size was 238

### Study procedures

Swabs were taken from inanimate surfaces and equipment and placed in Stuart transport media. Samples were immediately transported to the laboratory. They were cultured into MacConkey agar(Himedia Laboratories Pvt.Ltd, Nashik – India), Blood agar (Oxoid, Basingstoke Hampshire -United Kingdom (UK)) and Chocolate agar (Himedia Laboratories Pvt.Ltd, Nashik – India) and incubated at 35◦C -37◦C for 18 to 24 hours. The bacteria identification test was done using the Convectional method which involves Kligler Iron Agar(Oxoid, Basingstoke Hampshire -United Kingdom (UK)), Citrate(Oxoid, Basingstoke Hampshire -United Kingdom (UK)), Urea (Himedia Laboratories Pvt.Ltd, Nashik – India) and Sulphur Indole and Motility tests(Oxoid, Basingstoke Hampshire -United Kingdom (UK)).

### Data collection methods and tools

Data were collected by swabbing inanimate surfaces and equipment in pediatric wards by rubbing and rolling the wet and moistened swabs firmly several times across the sampling surfaces and equipment at noon hours (the time when hospital activities are already in progress). After rubbing and rolling, swabs were inserted into test tubes containing transport media (Stuart media) (Oxoid, Basingstoke Hampshire -United Kingdom (UK)), the common transport media used at KCMC. The samples reached the laboratory within 24 hours after collection for the culture process.

### Data Quality

Data collection sheets were checked for completeness and if correctly filled daily. Each procedure was performed according to the Standard Operating Procedures of the laboratory where the study was carried out. Organisms used for quality control were *Staphylococcus aureus* (ATCC-25923), *Escherichia coli* (ATCC-25922), and *Pseudomonas aeruginosa* (ATCC-27853).

### Data analysis

Data were cleaned and checked for completeness and consistency before analysis. Analysis was done by using SPSS version 20 (Norman H. Nie, Chicago, USA) and the results of proportion were presented in terms of tables.

### Ethical Consideration

Ethical approval for this study was obtained from the KCMC University Research and Ethics Review Committee (KURERC) with certificate number 11. Permission to conduct the study was obtained from the KCMC Hospital director. The informed consent form was not applicable since the study was dealing with only bacterial contamination of inanimate surfaces and equipment and not human participation.

## Results

A total of 238 swab specimens from inanimate surfaces and equipment resulted in 369 bacteria isolates. P1 samples were 71 (29.8%), P2 samples were 47 (19.8%), P3 samples were 60 (25.2%) and PICU samples were 60 (25.2%) (Table 01)

**Table 01:** Samples collected in each pediatric ward, n= 238.

| WARD | Frequency (n) | Percentage (%) |
| --- | --- | --- |
| Pediatric one (P1) | 71 | 29.8 |
| Pediatric two (P2) | 47 | 19.8 |
| Pediatric three (P3) | 60 | 25.2 |
| Pediatric ICU (PICU) | 60 | 25.2 |

### Proportion of contamination of inanimate surfaces and equipment

From the 206 (86.6%) samples out of 238, the bacterial isolates were 369. Gram-negative bacteria were the most dominant with 304 (82.4%) out of 369 positive isolates and Gram-positive were 65 (17.6%) out of 369 positive isolates. *E. coli* was the most leading isolate 114 (30.9%), followed by *Pseudomonas aeruginosa* 93 (25.2%), *Coagulase negative staphylococcus* 58 (15.7%), *Klebsiella oxytoca* 44 (11.9%), *Klebsiella pneumoniae* 41 (11.1%), *Acinetobacter* species 10 (2.7%), *S.aureus* 7 (1.9%), *Proteus vulgaris* 1 (0.3%) and *Citrobacter* species 1(0.3%) (Table 02).

**Table 02:**
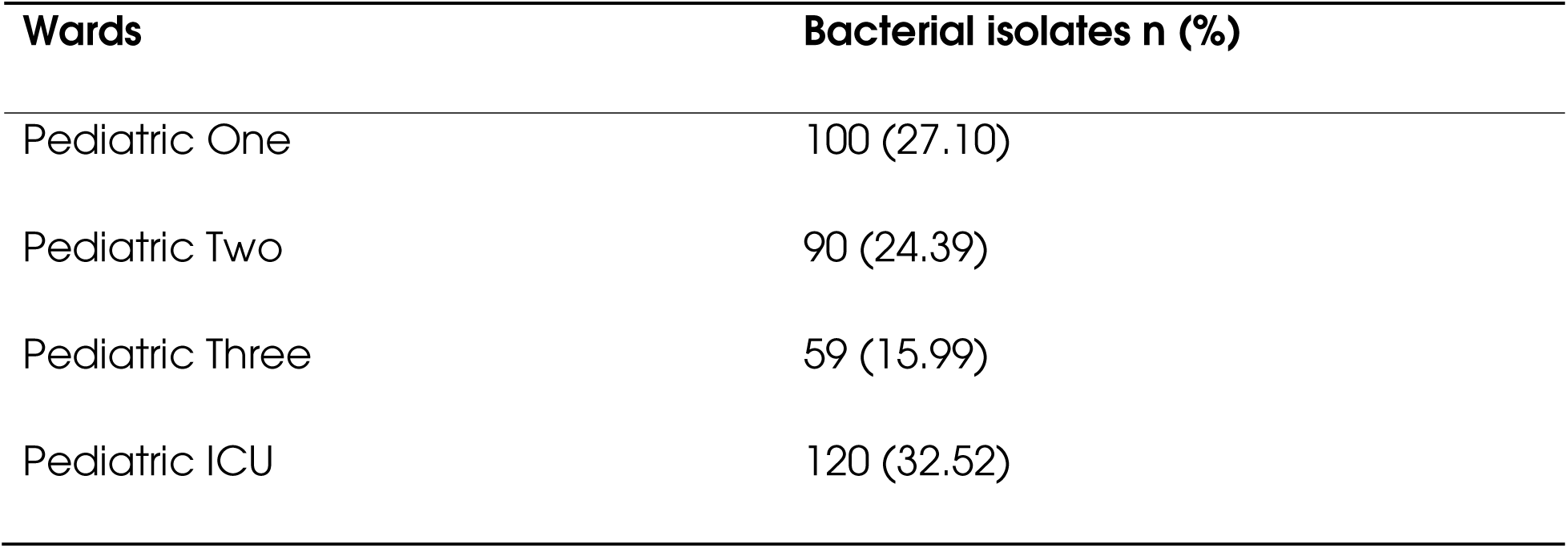
Distribution of bacteria isolated from each Pediatric ward, n=369.

### Types of bacterial isolates that contaminated inanimate surfaces and equipment

A total of 369 bacteria were isolated from sample swabbed on different surfaces and equipment in which ventilators found in pediatric ICU were highly contaminated 67 (18.2%) with different bacteria isolates, followed by washing sink tap water 52 (14.1%), stethoscope 44 (11.9%), door handles 36 (9.8%), electrical equipment 34 (9.2%), thermometers 34 (9.2%), medical charts 27 (7.3%), bedside tables 26 (7%), beds 25 (6.8%) and baby coats 24 (6.5%) (Table 03).

**Table 03:**
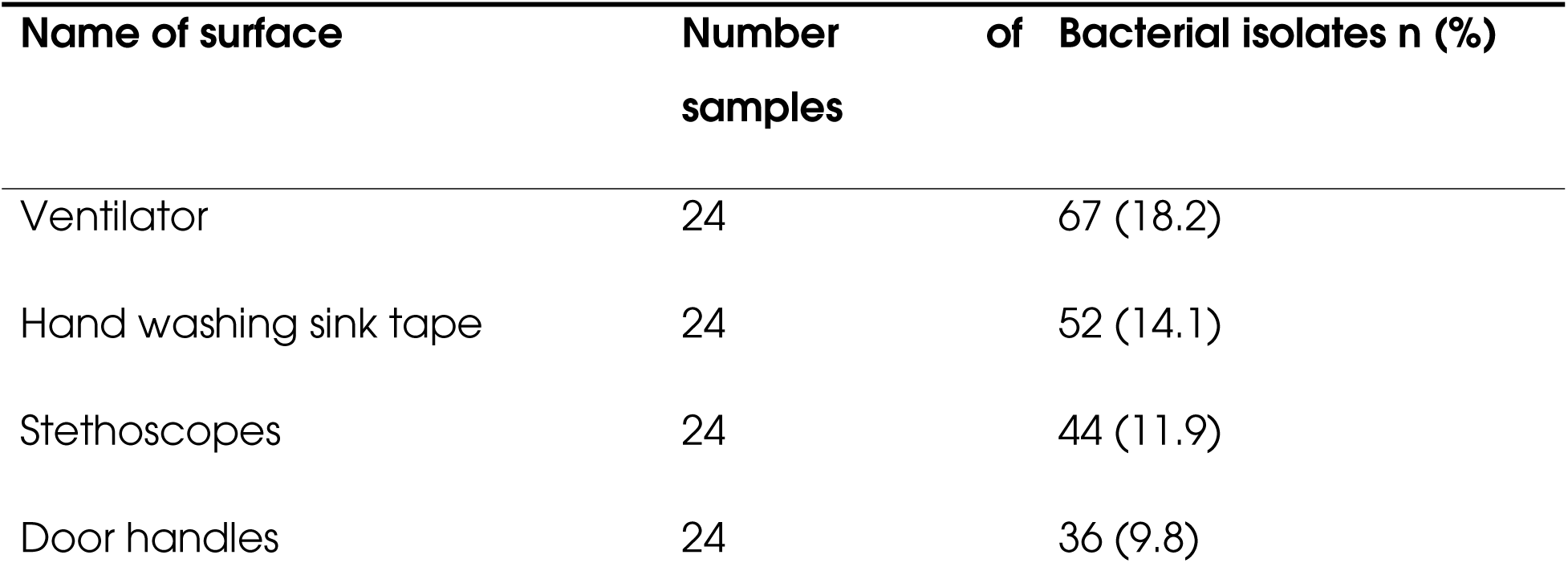
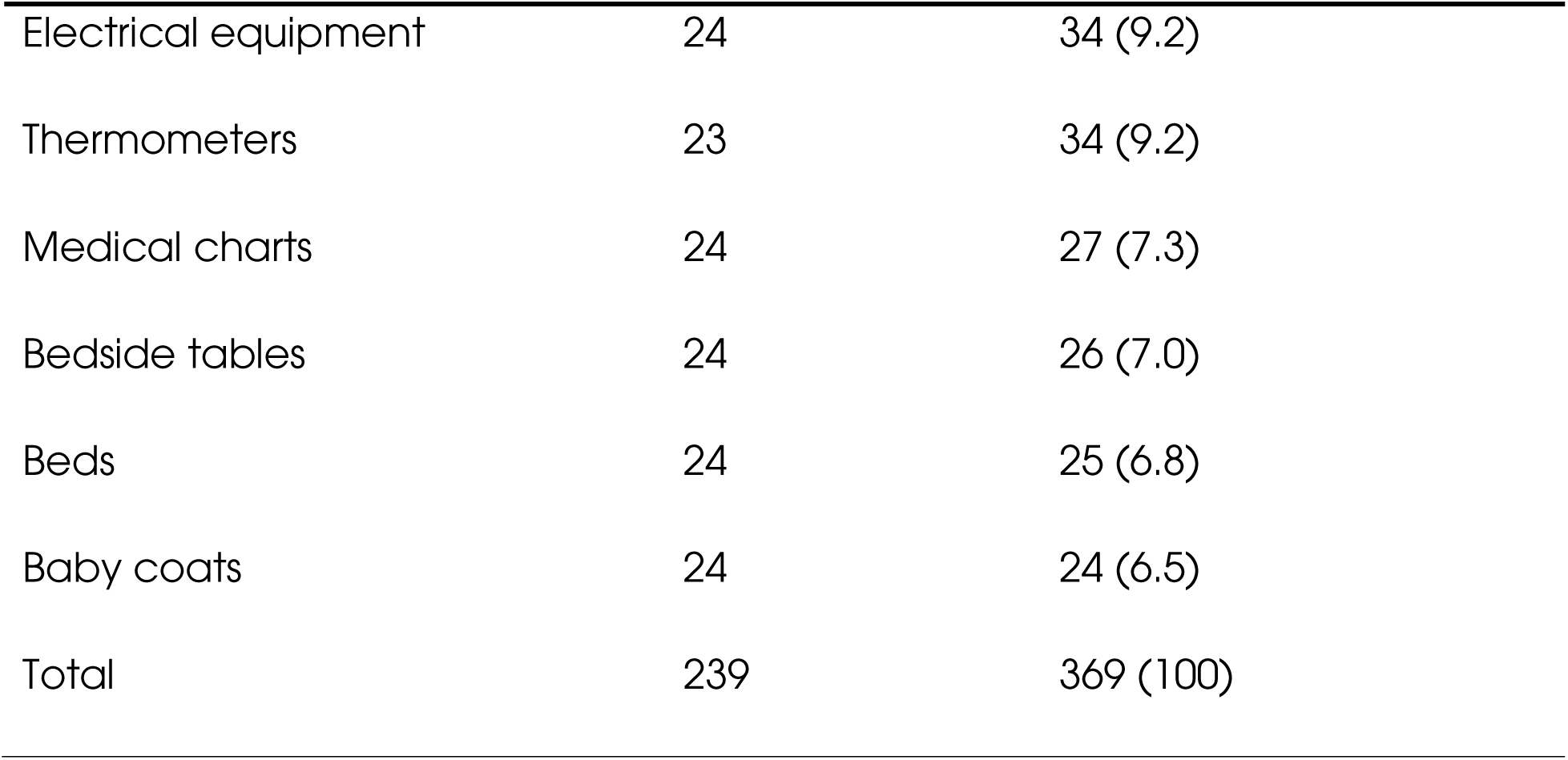
Total number of swabbed inanimate surfaces and equipment and types of bacterial isolates in pediatric wards at KCMC.

| Name of surface | Number of samples | Bacterial isolates n (%) |
| --- | --- | --- |
| Ventilator | 24 | 67 (18.2) |
| Hand washing sink tape | 24 | 52 (14.1) |
| Stethoscopes | 24 | 44 (11.9) |
| Door handles | 24 | 36 (9.8) |
| Electrical equipment | 24 | 34 (9.2) |
| Thermometers | 23 | 34 (9.2) |
| Medical charts | 24 | 27 (7.3) |
| Bedside tables | 24 | 26 (7.0) |
| Beds | 24 | 25 (6.8) |
| Baby coats | 24 | 24 (6.5) |
| Total | 239 | 369 (100) |

### Susceptibility pattern

The susceptibility test was performed for gram-negative and gram-positive bacteria based on the Kirby-Bauer disk diffusion method on Mueller-Hinton agar (MHA) (Oxoid, Basingstoke Hampshire -United Kingdom (UK)). All Gram-negative bacteria were resistant to Amox/clav and Gentamicin, and *S. aureus* was resistant to all Antibiotics except Ciprofloxacin (Table 04).

**Table 04:** Antimicrobial susceptibility test of gram-negative and positive bacteria from the inanimate surfaces and equipment in pediatric wards at KCMC. Resistance is represented by R while susceptible phenotypes are represented by S.

| ISOLATES | AMC | SXT | CIP | GN | MEM | AK | CAZ | C | TZP | VA | D |
| --- | --- | --- | --- | --- | --- | --- | --- | --- | --- | --- | --- |
| *S. aureus* | R | R | S | I | NT | NT | R | NT | NT | R | R |
| *P. aureginosa* | R | NT | NT | R | S | S | S | NT | R | NT | NT |
| *K. pneumonia* | R | R | S | I | S | NT | R | S | NT | NT | NT |
| *K. oxytoca* | R | R | S | R | S | NT | R | S | NT | NT | NT |
| *E. coli* | R | S | S | S | S | NT | S | S | NT | NT | NT |
| *Proteus vulgaris* | R | R | S | R | I | NT | R | R | NT | NT | NT |
| *Acinetobacter spp* | R | S | S | S | S | NT | S | I | NT | NT | NT |
| *Citrobacter spp* | R | R | R | R | R | NT | R | R | NT | NT | NT |

R- Resistance, S- Sensitive; I- Intermediate; NT- Not Tested
AMC- Amox/Clav, SXT- Trimethoprim/Sulfamethoxazole, CIP- Ciprofloxacin, GN- Gentamicin, MEM- Meropenem, AK- Amikacin, CAZ- Ceftazidime C- Chloramphenicol, VA- Vancomycin, D- Clindamycin

## Discussion

The observed overall proportion (86.6%) of bacterial contamination of inanimate surfaces and equipment in pediatric wards is significantly high, which poses a serious risk of nosocomial infections, as also observed from recent studies within East Africa and beyond ^24–27^. This elevated contamination rate could be attributed to inadequate adherence to Infection Prevention and Control (IPC) protocols, as proposed from surveillance studies across sub-Saharan Africa ^28,29^. Despite the intensified emphasis on IPC measures particularly during the COVID-19 pandemic, these findings highlights significant gaps in the consistent implementation of these measures.

*E. coli* isolates accounted for the most predominant bacteria followed by *Pseudomonas aeruginosa* in this work. The reason for the higher prevalence of *E. coli* and *P.aeruginosa* could be their wide adaptive capacity, which enables them to thrive in a wide range of conditions across environments and clinics ^30^.

On clinical surfaces, these pathogens exhibit a high capacity to form biofilms and resist the effects of antibiotics and disinfectants, thereby increasing the risk to hospitalized patients. *E.coli* is frequently associated with fecal contamination, which may result from inadequate surface cleaning or gaps in personal hygiene practices among healthcare workers. Similarly, *P.aeruginosa*, a waterborne pathogen, thrives in moist environments such as sinks and improperly sanitized medical equipment, underscoring deficiencies in surface decontamination procedures. These pathogens, alongside other isolates such as *coagulase-negative staphylococci, Klebsiella oxytoca, Klebsiella pneumoniae, Acinetobacter species, Staphylococcus aureus, Proteus vulgaris,* and *Citrobacter species*, poses a significant risk to vulnerable pediatric patients. The potential for severe infections is further exacerbated by increasing antimicrobial resistance, with many of these organisms emerging as common clinical isolates associated with multidrug-resistant nosocomial infections ^31^. While there is considerable information on the AMR pattern of *E. coli, K. pneumoniae* and other pathogens detected in this work, the AMR profile of *K. oxytoca* observed here suggests an overlooked threat, which has also been recently emphasized from comprehensive genomic analysis of AMR *K. oxytoca* by Maghembe et al^32^. This calls for serious studies on this category of pathogens in order to establish their clinical and public health risks, especially their AMR profiles. Similarly, with limited information on *Citrobacter* prevalence and AMR data, evidence is emerging pointing to *C. freundii* and *C. braakii* as clinically important pathogens with unprecedented AMR genotypes and phenotypes ^33^. Thus, findings from this study reiterate the need for attention to *Citrobacter* species locally and internationally.

The findings of this study indicate a bacterial contamination rate of 206 instances (86.6%), which aligns with the 86% reported in a study conducted at Addis Ababa Hospital in Ethiopia ^34^, However, this rate is higher compared to the 76% observed at Kishan University of Medical Science and Health Hospital in Iran ^5^, 66.7% in pediatric wards of Nigerian hospitals ^35^, 44.2% from Kawolo General Hospital in Uganda, ^22^, and 25.6% from referral hospitals in Assiut City, Egypt ^36^. These results demonstrate a significant level of bacterial contamination on inanimate surfaces and equipment in the pediatric wards of KCMC Hospital. The contamination rate found in this study is lower than the 88.5% reported in in Ayder Comprehensive Specialized Hospital, Mekelle, Ethiopia ^37^, which may be attributed to inadequate decontamination practices and substandard hand hygiene. A similar study conducted in Dodoma, Tanzania reported a bacterial contamination rate of 61.4% in which the leading isolate was *Staphylococcus aureus* ^38^

A notably high proportion of *Escherichia coli* was identified, with 114 instances (30.9%), in comparison to the 2.7% reported in Assiut City, Egypt ^36^ and 3.1% in Nigerian hospitals ^35^. Furthermore, a substantial proportion of *Pseudomonas aeruginosa* was recorded at 93 instances (25.2%), significantly higher than the 1.3% rate documented in pediatric wards of Nigerian hospitals ^35^. On the other hand, the proportion of *Staphylococcus aureus* was relatively low at 7 instances (1.9%), compared to 20% reported in Nepal and 75.4% in Uganda ^22^.

Antimicrobial resistance remains one of the most critical global health threats, associated with nearly 700,000 deaths annually, with projections estimating approximately 10 million deaths by 2050 ^22^. In this study, *Staphylococcus aureus* demonstrated resistance to Vancomycin; however, Vancomycin remains the most effective antibiotic against this pathogen. Additionally, resistance was observed among Gram-negative bacteria to Amoxicillin/Clavulanate, Trimethoprim/Sulfamethoxazole, Gentamicin, Ceftazidime, and Ciprofloxacin.

This may be attributed to the increasing overuse and misuse of antibiotics within the general population, exacerbated by a fragile healthcare management system that permits access to antibiotics without proper clinical prescriptions. The presence of resistant organisms such as *Klebsiella pneumoniae, Klebsiella oxytoca, Pseudomonas aeruginosa,* and *Escherichia coli* presents a considerable challenge to infection control and treatment outcomes. These findings are consistent with those from Kawolo General Hospital in Uganda ^22^ underscoring the urgent need for effective antimicrobial stewardship programs, routine surveillance of resistance patterns, and the improvement of infection control practices within healthcare settings.

In summary, the study highlights a critical burden of bacterial contamination on inanimate surfaces and equipment, particularly in pediatric wards. These findings reinforce the need for routine decontamination practices, enhanced infection prevention protocols and an antimicrobial stewardship program in Tanzania’s healthcare settings.

### Study Strengths and Limitations

Since our study is Hospital-based and focused solely on pediatric wards, the findings may not be generalizable to all Hospital settings at KCMC. However, the study provides valuable insights into bacterial contamination of inanimate surfaces and equipment, which may be relevant to other Hospital wards.

## Conclusion and Recommendation

The findings of this study revealed a significant burden of bacterial contamination and antimicrobial resistance in pediatric wards, emphasizing the urgent need for targeted interventions to reduce nosocomial infections and enhance patient safety. Our results identify a high proportion of pathogenic bacteria, including *E. coli, P.aeruginosa,* and *Klebsiella* species, as the primary contaminants. This suggests that bacterial contamination of surfaces and equipment is likely a major contributor to Hospital-acquired infections (HAIs). In pediatric wards, surfaces and equipment can harbor harmful bacteria that pose a particular risk to immunocompromised or critically ill patients. To mitigate this risk, it is essential to evaluate infection prevention practices, implement routine disinfection protocols, and raise awareness among healthcare professionals regarding the potential for pathogen transmission from hospital surfaces and equipment.

## Data Availability

All data produced in the present work are contained in the manuscript

## Acknowledgment

We extend our sincere gratitude to the administration of KCMC University and KCMC Hospital for granting permission to conduct this study. We also wish to express our deep appreciation to the School of Public Health at KCMC University for their valuable contributions and insightful suggestions during the planning and development of this research work. Their willingness to generously dedicate their time is greatly appreciated.

